# Upregulation of Reward Mesolimbic Activity via fMRI-Neurofeedback Improves Vaccination Efficiency in Humans

**DOI:** 10.1101/2024.09.18.24313899

**Authors:** Nitzan Lubianiker, Tamar Koren, Meshi Djerasi, Margarita Sirotkin, Neomi Singer, Itamar Jalon, Avigail Lerner, Roi Sar-el, Haggai Sharon, Hilla Azulay-Debby, Asya Rolls, Talma Hendler

**Affiliations:** School of Psychological Sciences, Tel Aviv University, Tel Aviv, Israel; Department of Psychology, Yale University, New Haven, CT, USA; Princeton Neuroscience Institute, Princeton University, Princeton, NJ, USA; Department of Immunology, Rappaport Faculty of Medicine, Technion - Israel Institute of Technology, Haifa, Israel; Department of Neuroscience, Rappaport Faculty of Medicine, Technion - Israel Institute of Technology, Haifa, Israel; Sagol Brain Institute, Wohl Institute for Advanced Imaging, Tel Aviv Sourasky Medical Center, Tel Aviv, Israel; Sagol School of Neuroscience, Tel Aviv University, Tel Aviv, Israel; Montreal Neurological Institute, McGill University, Montréal, QC, Canada; Sackler Faculty of Medicine, Tel Aviv University, Tel Aviv, Israel

## Abstract

The placebo response is associated with a positive expectation of recovery. Recent studies in mice uncovered a potential underlying mechanism of placebo effects, by demonstrating the involvement of the dopaminergic mesolimbic pathway, previously implicated in reward expectation, in immune modulation. Yet, it is not known whether an analogous brain-immune regulatory mechanism exists in humans, and whether it employs conscious positive expectations. Here, we employed fMRI-neurofeedback to train healthy participants to increase their reward mesolimbic activity by using self-chosen mental strategies, followed by Hepatitis B Virus (HBV) vaccination. We found that stronger Ventral Tegmental Area (VTA) activity, but not control regions activation, was associated with higher post-vaccination HBV antibody titer. Interestingly, higher VTA activity before vaccination was associated with mental strategies characterized by positive expectation. Thus, our results establish a relationship between reward system activity, positive expectation and immunity in humans and reveal a potential neuropsychological mechanism for non-invasive immune modulation.

https://clinicaltrials.gov/study/NCT03951870.

## Introduction

Could our thoughts and feelings directly affect our physical well-being? While clinical benefits of positive expectations from medical treatment (i.e. placebo effects) have been extensively documented^1^, the underlying neural mechanism of such a mind-body link has only begun to be elucidated. The neural network associated with reward expectation, exploration and prediction error is the dopaminergic mesolimbic pathway, specifically the Ventral Tegmental Area (VTA)^2^. Previous studies in mice demonstrated that chemogenetic activation of dopaminergic neurons in the VTA alters immune activity, which results in improved recovery from bacterial infection^3^ and attenuated lung tumor growth^4^. Using optogenetic stimulation, it was further shown that phasic activation of the VTA increases immune activity. Intriguingly, a prototypical increase in mating-induced cytokines was abolished by specific inhibition of VTA dopaminergic activity^5^. These findings demonstrate the top-down regulation of peripheral immunity in the context of reward-seeking behaviour. In humans, correlations between reward-related neural activity (measured through functional neuroimaging) and specific immune markers have been documented^6–8^. However, it is unclear to what extent the human reward mesolimbic (reward-ML) pathway causally modulates immune function, and if it does, through which psychological processes.

In the current study we investigated whether upregulation of the reward-ML network (the VTA and bilateral nucleus accumbens (Nac); see **Figure S1**) can enhance the response to an immune challenge. We hypothesized that upregulation of reward-ML activity could enhance participants’ immune response to vaccination (i.e., antibody titer). To test this, we applied functional Magnetic Resonance Imaging NeuroFeedback (fMRI-NF) training^9–11^, a self-neuromodulation procedure based on reinforcement learning principles. In each fMRI-NF trial (**Figure 1b**), participants chose and applied a mental strategy – a combination of perceptual, affective, cognitive, or meta-cognitive mental contents – and then received reinforcing feedback reporting on changes in targeted brain regions activations, thus learning which mental strategies induce the desired upregulation pattern. These self-chosen strategies were recorded during NF training, and then systematically characterized (**Figure 1c**, upper panel, and **Figure S3**).

**Figure 1:**
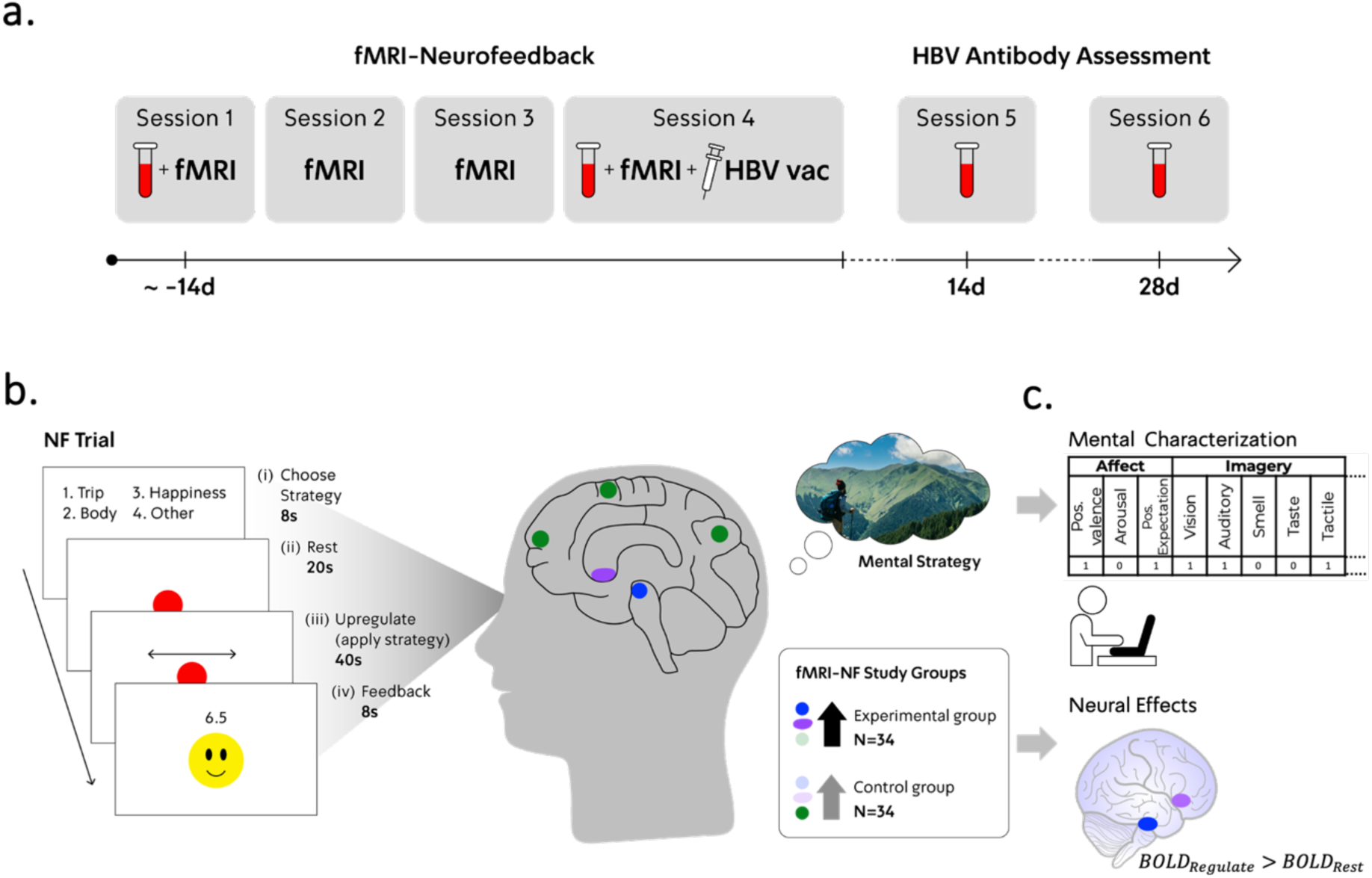
**a. Experimental procedure:** NF participants performed 8 to 11 fMRI-NF runs (each run consists of five consecutive trials) across three fMRI-NF sessions (session 2 to 4), or, if time allowed, four fMRI-NF sessions (session 1 to 4). Session 4 fMRI-NF was immediately followed by HBV vaccination. Blood tests were acquired to assess HBV antibody titer (baseline at sessions 1 and 4; post-vaccination measurements at sessions 5 and 6). **b. NF procedure:** During each NF trial, participants were instructed to (i) choose via a key press one out of four mental strategies (predefined by participants before each NF run) for the upcoming ‘Upregulate’ screen, (ii) rest while passively watching the red dot on the screen, (iii) upregulate neural target activity by employing their chosen strategy (for example, recalling a trip), and (iv) receive numeric (1-10) and graphic (smiley) feedback proportional to regulation success (difference in neural target activity between the current trial’s ‘Upregulate’ and ‘Rest’ epochs). The experimental group received feedback contingent on the reward-ML network (VTA (blue) and bilateral Nac (purple)). Control NF group participants were randomly assigned to receive feedback contingent on one of four functional networks unrelated to reward processes (an example network is illustrated in green; see ‘Online Methods’ and **Figure S2**). Guided by repeated feedback, participants learn which strategy types are the most effective for facilitating the desired upregulation effect. **c. Neuropsychological analyses of NF data.** Upper panel: Mental characterization included post-session labelling of self-chosen mental strategies across 45 mental features, resulting in a vectorial mental characterization of each NF trial. Lower panel: Neural effects were estimated based on reward-ML Regions Of Interest (ROI) analyses of differences between ‘Upregulate’ and ‘Rest’ fMRI-BOLD activity, across NF training runs.

## Results

In a pre-registered, double-blind, randomized controlled trial, 85 healthy individuals were recruited to the study (18-45, M_age_±SD: 24.98 ± 4.56 years, 51 females). Participants were assigned either to upregulate the joint activity of their reward-ML network (experimental group; N=34), to upregulate one out of four non-mesolimbic functional networks (control group; N=34) (**Figure 1b**), or to a no-NF group (N=17) (see **Table-1**). Immediately after the last training session/waiting period, all participants received a Hepatitis B Virus (HBV) vaccine to challenge their immune system. Blood samples were drawn twice before and twice (14 and 28 days) after vaccination to assess the development of HBV antibodies (**Figure 1a** and ‘Online Methods’). A subgroup of subjects (n=34) arrived for a 7^th^ session for a blood test 3 months following vaccination, aimed to assess long-term adaptive immune system effects manifested by HBV antibodies. The main trial began in March, 2020, and ended in August, 2022, when reaching the pre-registered N. No harms or unintended effects occurred to participants during the study.

**Table 1:** Demographics and baseline motivational and personality characteristics.

p- values were calculated based on a one-way ANOVA across the three study groups.
| Variable | Reward-ML NF | Control NF | no NF | p-value |
| --- | --- | --- | --- | --- |
| N | 34 | 34 | 17 | N/A |
| Gender (F/M) | (21/13) | (21/13) | (9/8) | N/A |
| Age | 24.53 (4.43) | 24.16 (2.81) | 27.67 (6.66) | 0.036 |
| TPQ Novelty Seeking | 14.96 (4.03) | 14.84 (3.91) | 14.2 (6.09) | 0.889 |
| TPQ Harm Avoidance | 10.75 (5.69) | 10.71 (5.59) | 11.4 (5.85) | 0.941 |
| TPQ Reward Dependence | 21.24 (2.57) | 20.49 (3.28) | 19.0 (3.33) | 0.138 |
| NEO-FFI Neuroticism | 28.11 (7.79) | 30.26 (7.72) | 29.4 (8.71) | 0.581 |
| NEO-FFI Openness | 38.64 (6.49) | 41.5 (5.54) | 42.7 (7.12) | 0.107 |
| NEO-FFI Agreeableness | 52.07 (6.28) | 50.58 (6.12) | 47.5 (8.02) | 0.165 |
| NEO-FFI Conscientiousness | 47.75 (5.78) | 47.28 (7.05) | 45.9 (4.79) | 0.727 |
| NEO-FFI Extraversion | 43.01 (6.32) | 42.39 (8.12) | 43.0 (8.15) | 0.942 |
| SPSRQ Reward Sensitivity | 9.68 (3.82) | 10.48 (3.48) | 9.8 (4.58) | 0.698 |
| SPSRQ Punishment Sensitivity | 10.21 (4.43) | 9.97 (4.47) | 9.5 (5.15) | 0.912 |

### Neurofeedback Regulation Effects

We first determined whether the experimental group would exhibit elevated reward-ML activation during NF training compared to the control NF group (our neural primary outcome measure). For each mesolimbic region, we conducted a mixed-linear-effects regression analysis. The dependent variable was Blood-Oxygenated-Level-Dependent (BOLD) percent signal changes (PSC) of ‘Upregulate>Rest’ contrasts calculated per NF run (five consecutive NF trials). Hypotheses were quantified through group and time (session 2 to 4; see ‘Methods’ section) terms and group-by-time interaction term as fixed effects. A nested runs-within-sessions term was introduced as a fixed effect to account for within-session fatigue, and random intercepts were introduced to account for individual variability. Analyses revealed that VTA BOLD activity significantly increased across sessions for all NF participants (VTA main time effect: F(2,66)=7.61, p<0.001, *η*^2^*_partial_* = 0.19, **Figure 2a**, upper panel), but with no significantly larger increases in the reward-ML group compared to the control group (VTA group-by-time interaction: F(2,66)=0.762, p=n.s). Nac activity marginally increased more in the reward-ML group than in the control group (Nac group-by-time interaction: F(2,66)=2.708, p=0.067, *η*^2^*_partial_* = 0.077; session-4 simple group effects: t(66)=2.42, p=0.019, Cohen’s D=0.295; **Figure 2b**, lower panel). Together, these results indicate differential effects across the reward-ML network, with substantial VTA upregulation effects regardless of NF targeting.

**Figure 2:**
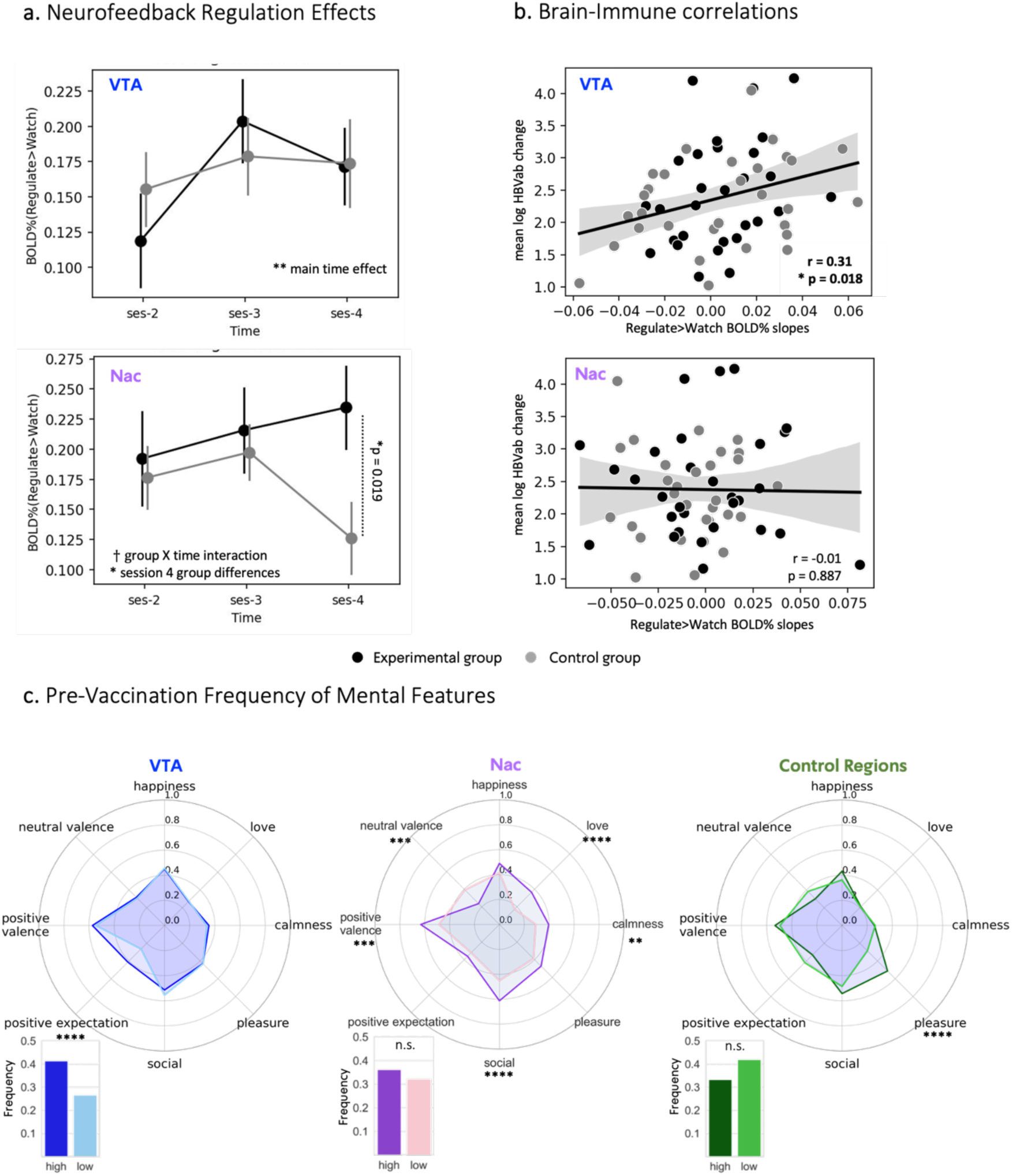
**a. Neurofeedback regulation effects.** fMRI-NF regulation effects per group (Mean±SE; black - experimental group; grey - control NF group) for VTA (upper panel; blue) and Nac (lower panel; purple) **b. Brain-Immune correlations.** Pearson’s correlations between the degree of regulation in each region (x-axes; higher positive slopes indicate a more substantial increase in regional activity across NF training runs), and post-vaccination change in HBV antibody levels (y-axes; higher values indicate a more substantial increase in HBV antibody levels). **c. Pre-vaccination frequency of mental features.** Frequency of affective and reward-related features in mental strategies during session 4 (prior to vaccination), for high and low regional activity levels in the VTA, Nac and non-mesolimbic control regions (coloured by dark vs bright shades, respectively). † p<0.1; * p<0.05; ** p<0.01; *** p<0.001; **** p<0.0001.

### Associations between reward mesolimbic upregulation and vaccination efficiency

The VTA upregulation effects indicate that both experimental and control NF groups experienced reward-related activations. Inspecting NF groups separately via mixed-linear-effects regression analysis (similar to the model detailed above, excluding group-related terms) revealed significant increases across sessions in VTA activity in the experimental group (F(2,33)=5.02, p<0.001, *η*^2^*_partial_* = 0.233) and a marginal increase in the control NF group (F(2,33)=2.66, p=0.071, *η*^2^*_partial_* = 0.139). Indeed, NF training is a reward learning paradigm (see also **Figure S4** demonstrating reward-related activations). In line with these results, we hypothesized that regardless of NF group allocation, reward-ML activity during NF training would be correlated with an elevated post-vaccination immune response.

We assessed the correlations between neural regulation effects and immune responses (our first pre-registered hypothesis for the primary immunological outcome measure). For each region, participant-specific regulation effects were quantified as the individual change (i.e. increases or decreases, captured by a linear slope; see refs^12–14)^ in ‘Upregulate>Rest’ BOLD activity levels across NF runs. IgG/IgM HBV antibody (HBVab) levels in blood plasma were log-transformed. The immune response was measured as the difference between mean post-vaccination (sessions 5 and 6) and mean baseline (sessions 1 and 4) measurements. Seven vaccination non-responders (post-vaccination HBVab levels<10 mg/ml; a known phenomenon specifically for HBV vaccination, see ref^15^), evenly distributed among NF groups, were excluded from further immunological analyses. One participant dropped out due to personal reasons before post-vaccination antibody assessment sessions, leaving 60 participants for analyses (30 in each NF group). Correlational analyses revealed that stronger increases in regulation of VTA, but not of Nac, were significantly correlated with higher post-vaccination HBV antibody changes (VTA: r=0.31, N=60, p=0.018, *CI*_95%_= [0.061, 0.523]; **Figure 2a**, right panel; Nac: r=-0.01, N=60, p=0.887, *CI*_95%_= [-0.263, 0.244]; **Figure 2b**, right panel; Bonferroni-corrected critical p=0.05/2 = 0.025). These findings suggest the existence of a VTA-immune modulatory effect in humans, which corresponds with previous findings in mice^3–5^.

To determine whether these brain-immune associations persisted in time, we examined correlations between reward-ML regulation effects and HBVab change three months following HBV vaccination (secondary immunological outcome measure). Out of 34 participants with long-term blood test measurements, 27 were NF participants. Three non-responders (according to the threshold defined above) were excluded, leaving 24 participants for analysis (12 per NF group). Correlational analyses revealed that the pattern of results exhibited two weeks to one month after vaccination was preserved three months following vaccination (VTA: r=0.3, p = 0.152, *CI*_95%_= [-0.116, 0.628]; Nac: r=-0.19, p = 0.365, *CI*_95%_= [-0.551, 0.231]), although it did not reach significance, possibly due to low statistical power.

Critically, an alternative, non-causal explanation for the observed correlation between VTA regulation and HBVab change could be that general success in the NF task, rather than specific VTA regulation, led to a stronger immune response (at least for participants from the experimental group whose rewarding feedback was partly contingent on VTA levels). To test this possibility, we examined whether activity in the non-mesolimbic control regions, reinforced during regulation for the control group participants, was correlated with HBVab change. However, our analyses did not reveal such a correlation (r=0.11, N=30, p=0.559, *CI*_95%_= [-0.26, 0.452]). In addition, VTA or Nac activity during session-4 ‘Feedback’ condition (Figure 1b), which captures neural reward responsivity levels during NF training, did not correlate with HBVab change (VTA: r=0.15, p=0.238, *CI*_95%_= [-0.11, 0.389]; Nac: r=0.04, p=0.763, *CI*_95%_= [-0.216, 0.291]). Together, these analyses rule out general NF learning or reward effects as the driving factor of a VTA-immune correlation.

Another non-causal explanation for the VTA-immune correlation is that individuals with higher trait motivational capacities may exhibit both stronger VTA regulation effects and higher vaccination efficiency, regardless of the temporal contingency between NF training and HBV vaccination. To examine this possibility, we assessed whether post-vaccination HBVab change and VTA upregulation effects were correlated with an array of motivational neuro-behavioural trait indices assessed by designated tasks and questionnaires collected before NF training (pre-registered as secondary outcome measures). These included fMRI BOLD responses to rewards in a Monetary Incentive Delay task^16^, a behavioural measure of effort expenditure levels^17^, and approach-avoidance tendencies^18^. However, analyses did not reveal any such associations (see ‘Supplementary Information’), ruling out such individual differences in motivational traits as the cause of the VTA-Immune correlation.

We next examined our second pre-registered hypothesis regarding the primary immunological outcome measure, that the experimental group would exhibit a larger increase in anti-HBV antibody titer compared to the control group and the no-NF group. A one-way ANOVA across the three study groups (N=72 after excluding vaccination non-responders, as specified earlier) with post-vaccination HBVab change as the dependent variable did not reveal significant differences between groups (F(2,70) = 0.65, p = 0.565). This is contrary to our hypothesis but in line with the lack of significant differences between NF groups in VTA regulation effects, which were found to be correlated with, and possibly drive, post-vaccination HBVab change.

### Specificity of the link between VTA upregulation and positive expectation

To reveal the psychological aspects of the link between VTA upregulation and immune function, we tested whether elevated reward-ML activity before HBV vaccination (during session 4) was associated with the employment of mental strategies characterized by reward-related features (pre-registered as a secondary outcome measure), and specifically positive expectation, which was previously reported to precede placebo effects^19,20^. As for brain-immune correlations, we analyzed data from both NF groups together. These analyses were defined as exploratory.

To quantify the frequency of mental features during session 4 fMRI-NF training, we employed a new methodology for a trial-locked estimation of mental strategies during regulation attempts. For each NF trial, participants chose via key press which strategy to apply (**Figure 1b**, choice screen). Following training, participants characterized their applied strategies via a dedicated Mental Strategies Questionnaire for NF (**Figure S3**). This resulted in a binary matrix that indicates the involvement of mental categories/dimensions during each NF trial.

For each target region, we classified the fourth NF session of each participant (*N_subject_* = 68) as exhibiting either high or low activity patterns (based on a median split of session 4 ‘Upregulate>Rest’ BOLD activity levels). We then compared the frequency of mental features in trials from sessions exhibiting high vs low activity, using Chi-squared tests (29 included features, *p_Bonferroni_* = 0.05/29 = 0.0017; total *N_trails_* = 962. Feature-specific N differed according to participants’ responses (trials in which participants did not classify a strategy for a certain feature were excluded), and is reported below for each Chi-square test; see more details in ‘Online Methods’). Focusing on reward-related and affective features, we found that elevated VTA activity was associated solely with mental strategies characterized by a higher frequency of positive expectation feature (‘*Experiencing a positive expectation or excitement towards events/occurrences*’) (χ²(1)=17.33, p<.0001, *N_trails_* = 862, *φ* = 0.140), while other reward-related/affective features did not differ as a function of VTA activity (**Figure 2c**, left panel). Conversely, elevated Nac activity was associated with higher frequency of several reward-related/affective features, such as love (χ²(1)=43.66, p<.0001, N = 962, *φ* = 0.213), social content (χ²(1)=24.40, p<.0001, *N_trails_* = 960, *φ* = 0.159), positive valence (χ²(1)=20.63, p<.0001, *N_trails_* = 962, *φ* = 0.146) and calmness (χ²(1)=10.95, p<.001, *N_trails_* = 962, *φ* = 0.107), but not with positive expectation (**Figure 2c**, middle panel). Similarly, elevated activity in non-mesolimbic regions regulated by participants in the control group (thus tested only on participants in the control group, N=34), was associated only with the pleasure feature (χ²(1)=16.49, p<.0001, *N_trails_* = 415, *φ* = 0.199; **Figure 2c**, middle panel), with no association to positive expectation, further supporting the specificity of the link between positive expectation and VTA upregulation in the context of NF training (see **Figure 2c**, right panel, and ‘Supplementary Information’ for complementary analyses). Taken together, these results suggest that high VTA upregulation before vaccination, but not Nac or control regions, was associated with a key antecedent of placebo responses – positive expectations.

## Discussion

Overall, this study demonstrates that upregulating the VTA with fMRI-NF training is associated with a stronger post-vaccination immune response in humans. By empirically ruling out central non-causal interpretations, our findings suggest a top-down brain-immune regulation mechanism, similar to that previously described in animals^3–5^, as a potential underlying neuropsychological mechanism of the placebo phenomenon. Utilizing a new methodology for characterizing mental strategies, we further support this claim by linking pre-vaccination VTA activation with positive expectations, which were shown to precede placebo effects^19,20^.

Of note, as the observed neuroimmune effects were isolated to the VTA, it is possible that these could have been amplified and even result in the hypothesized group differences if the experimental group neural target for up regulation was exclusively the VTA rather than the entire reward-ML network. Moreover, the association between VTA upregulation and mental strategies characterized by positive expectation suggests that future clinical studies could elicit stronger downstream immune effects by encouraging participants to employ mental strategies that evoke positive expectations. Of note, the current study was designated as a mechanistic investigation of the link between the reward-ML network and immune function in humans. Therefore, although we present a potential neural intervention for boosting vaccination efficiency in humans, the number of participants in our study is considered relatively small for testing the efficacy of an intervention in a clinical trial. In this regard, to better understand the extent of mesolimbic-immune regulation, it would be beneficial to conduct a larger, phase-2 clinical trial, and to investigate additional immune outcomes (e.g., cytokines, cellular profiling) and challenges (e.g., bacterial infection, inflammation and malignancy).

Altogether, our findings emphasize the “double life” of the reward mesolimbic pathway^21^, namely, guiding reward-seeking behaviour by anticipating external positive outcomes, while simultaneously boosting somatic resilience to better deal with the challenges that accompany the pursuit of our appetitive goals. Arguably, our self-neuromodulation fMRI-NF approach may be used to discover new ways to boost immune processes, with the potential for major therapeutic implications, in fields such as cancer immunotherapy and chronic inflammation. Thus, we may harness the natural capacities of our mind and brain to heal our bodies in times of need.

## Acknowledgements

This study was funded by Joy Ventures foundation and by KAMIN program in the Israeli ministry of innovation. We thank Yoav Benjamini for his statistical advice, Noam Noy for her assistance in creating the graphical illustrations depicted in Figure 1, and Areen Kuzli and Moran Szwarcwort for their assistance in measuring plasma antibody titer. N.L. would like to thank the Teva Bio-Innovation Forum.

## Authors contributions

N.L., T.K., A.R. and T.H. conceptualized the study and designed the experiments. N.L. and T.H conceptualized the MSQ questionnaire. N.L. developed the randomized-network control condition and the mental strategies characterization protocol. I.J. and N.S. developed online fMRI-NF analysis pipelines. N.L. and M.D. collected the behavioural, neural, and mental data. A.L., R.S. and H.S. collected blood samples and vaccinated participants. N.L. and M.D. analyzed the behavioural and neuroimaging data. T.K., R.S. and H.A.D collected and analyzed the immunological data. A.R. and T.H. secured funding and supervised the study. N.L., T.K., A.R. and T.H. wrote the paper. All authors edited the paper.

## Competing interests

T.H. is the Chief Medical Scientist and Chair of advisory board in GrayMatters Health Co Haifa Israel. T.H., A.R., N.L. and T.K. have a filed patent related to the topic of this paper in the United States Patent and Trademark Office (application number: 17/435.906; Title of Invention: Neurofeedback and Induction of an Immune Response). The other authors report no competing interests.

## Online Methods

### Pre-registration

Pre-registration can be found at the clinicaltrials.gov website, study ID: NCT03951870 https://clinicaltrials.gov/study/NCT03951870.

### Power calculation

To achieve sufficient statistical power (80%) for our study design, the various expected effects were analysed separately. Results indicated that the most demanding hypothesis, the existence of a moderate correlation (r=0.3) between reward mesolimbic activations during NF training and immunological outcome measures, requires at least 67 data points. As this correlation could be assessed based on subjects from both NF groups, NF group sizes were set to 34 each (with 17 subjects in the no-NF control group, resulting in a 2:2:1 ratio design).

### Participants

In total, 85 healthy individuals (ages 18-45, *M*_age_±*SD*: 24.98 ± 4.56 years, 51 females) who intended to receive vaccination for HBV were recruited for the study. Six additional participants were recruited to the study but dropped out either after the first session from personal reasons (N=5), or after their third session due to COVID pandemic restrictions (N=1), and were not included in analyses. Inclusion criteria to the study were normal or corrected to normal vision and being able to undergo MRI scanning. Participants were screened according to several exclusion criteria: autoimmune or other acute or chronic disease, previous diagnosis of psychiatric or neurologic disorders that led to hospitalization, chronic use of drugs, and MRI eligibility. An additional exclusion criterion was previous HBV vaccination (other than during infancy) to minimize initial HBV antibody levels, which is different than the pre-registered criterion of prior HBV vaccination in the last ten years. This criterion was adjusted based on pilot studies that revealed high baseline HBV antibody levels even for subjects that were vaccinated approximately 10 years prior to recruitment (see “Supplementary Information”). All participants received a detailed explanation of the study (without exposing them to any hypotheses) and provided informed consent according to the Tel-Aviv Sourasky Medical Center Institutional Review Board (IRB) committee guidelines, prior to participation. Subjects were compensated at a rate of 50 NIS (∼ $15) per hour.

Pilot studies aimed at assessing the immune response in our target population and the efficacy of our fMRI-NF training procedure were conducted between January and December 2019 (see “Supplementary Information”).

### General Procedure

All experimental sessions and data collection were conducted at the Tel-Aviv Sourasky Medical Center. Subjects were randomly assigned to one of three study groups: reward ML-NF (N=34), control-NF (N=34), or a no-NF control group (N=17). Allocation to NF groups was double-blinded throughout the whole experiment (see more details in ‘Randomization and double-blinding’). At the beginning of session 1, subjects were informed of the study goals and experimental protocol, and filled out an informed consent according to IRB guidelines. NF Subjects took part in four fMRI scanning sessions. Immediately prior to the first session, subjects filled out TPQ^1^, NEO-FFI^2^ and SPSRQ^3^ questionnaires, and performed the Effort Expenditure for Rewards Task (EEfRT) aimed to assess effort expenditure motivational tendencies^4^. Then, subjects received instructions about the NF task. To enable blinding, all subjects received NF task instructions appropriate to all targets of the study (see ‘NF double-blinding and randomization’ section). In general, subjects were told that to up-regulate (increase) neural activity during training, they will have to use mental strategies that could have several different characteristics, and that each region in the brain may be regulated with its own unique type of mental contents, which they will have to discover by themselves. It was also stressed that for each subject, different strategies may be efficient. Examples of mental strategies covered a variety of general categories: sensory, imagery or memory, emotional or motivational contents, social aspects, or conceptual/arithmetic. Other general instructions were to remain with one strategy per cycle (not to switch in the middle of a cycle), to make the strategy tangible, and relate as much as possible to the mental state they wish to employ, and during Rest screens to relax (but with eyes open), while making sure they do not engage in strategy application or choice.

The first fMRI scan lasted for approximately 90 minutes. A head localizer sequence and a structural scan were acquired. Following structural scans, subjects proceeded to an eight-minute functional resting-state scan (not analysed or reported in this paper). Then, subjects performed two runs of a functional localizer - the Monetary Incentive Delay (MID) task (details below). Individual mesolimbic ROIs were extracted algorithmically without hampering double-blinding, according to the subject’s group allocation (details below). If time allowed, subjects proceeded to either one or two NF task runs, each run consisting of five NF cycles (**Figure 1b**), and If not, participants started their NF training in the following session. During sessions 2 to 4, a head localizer sequence, a structural scan, and a functional EPI template were acquired. Then, subjects proceeded to three or four NF runs. At the fourth session, a post-NF training resting-state scan was ultimately obtained.

Upon finishing the last NF practice session, participants received the HBV vaccination at the Tel Aviv Sourasky Medical Center (Ichilov Hospital). To assess immunological outcome measures, all participants provided four blood samples to assess HBV antibody levels, before NF training (session 1), at the beginning of session 4, and two and four weeks following HBV vaccination (sessions 5 and 6, respectively).

Participants allocated to the no-NF group participated in the procedure described above except NF training. Specifically, in session 1 they filled out questionnaires, performed EEfRT, and participated in an fMRI scanning session with structural, resting state and MID scans only. In session 4 they only provided a blood sample and received HBV vaccination. Sessions 5 and 6 were identical to those of NF participants.

Finally, a subset of 34 participants (14 reward-ML NF, 13 control NF, and 7 no-NF) were randomly allocated to arrive at session 7 (not shown in Figure 1a) to provide an additional long-term blood test. Of note, a subset of 30 participants were planned to arrive to that follow-up session. However, several participants that were allocated to this group did not wish to attend the follow-up session. Thus, to reach the N=30 goal, we adapted the long-term assessment recruitment procedure by inviting participants according to availability and regardless of group allocation (which remained double-blinded). This solution led to an unbalanced representation of the no-NF group (consisting of 3 out of the 30 participants). Thus, to maintain a planned 2:2:1 ratio between groups as for the main immunological outcome measure, we invited 4 additional no-NF participants that were able to attend the three months follow up session, reaching the following Ns per group: reward-ML NF = 14; control NF = 13; and no-NF = 7. Importantly, these additional four subjects have no bearing on the brain-immune correlation reported in the manuscript (as no-NF subjects are not included in this analysis).

### Tasks

#### Monetary Incentive Delay (MID)

To assess the relationship between neural reward responsivity measures (reward anticipation and consumption) and implicit regulation of immune functions, we used a variant of the Monetary Incentive Delay (MID) task^5^ that was developed by Kirschner and colleagues (see refs^6,7^). This variant enables the assessment of reward anticipation and reward outcome across three reward levels, and uniquely enables parametric modulation of the consumption stage, which is calibrated according to individual task performance. The task was performed during session 1 fMRI scanning by all participants. Participants were informed that at the end of each of the two experimental runs, the software would sum the winnings of three randomly selected trials, and the amount of money they would win will appear on the screen. These sums were added to their compensation for participating in the experiment. The maximum amount of money to be won was 30 NIS.

During each trial, one of three different cues was presented for 0.75 s. The cue indicated the maximum possible amount participants could gain in that trial (i.e., 10 NIS, 2 NIS, or 0 NIS; 1 NIS = ∼$0.35 USD). After a delay of 2.5–3 s, the participants had to identify an outlier from 3 presented circles and press a button (a difference in either left or right circles, while the central circle was always constant) as rapidly as possible. Circles appeared for 0.32 to 1 s. Immediately following key-press, participants were notified of the amount of money they had won (duration of feedback: 2 s). The actual amount of money won for each trial was calibrated on the basis of the response times of the previous 15 individual trials, in order to account for individual differences in response time, and thus ensure similar frequencies of high rewards across individuals (roughly one-third for each level – high, moderate and low amounts). Error trials were defined as trials with a wrong response or a late response (more than 1 s) and were excluded from analyses.

Each participant performed two training runs, one outside and another inside the scanner. Excluding the training sessions, the task included two experimental runs with 36 trials of about 10 s each. The intertrial interval (ITI) was jittered from 1 to 9 s with a mean of 3.5 s to enhance statistical power. In total, a run lasted 6:32 minutes. The task was implemented using the MATLAB toolboxes Cogent 2000 and Cogent Graphics.

#### Effort Expenditure for Rewards Task (EEfRT)

To assess the relationship between incentive salience and immune function, all subjects performed the EEfRT^4,8^ at the beginning of the first experimental session outside the scanner.

In the EEfRT, participants choose between two different task difficulty levels on each trial to obtain monetary rewards. For both types of trials, participants were asked to perform repeated manual button presses within a short period, where each button press raises the level of a virtual “bar” viewed onscreen by the participant. Participants are eligible to win the money allotted for each trial if they raised the bar to the “top” within the prescribed time period. Successful completion of hard-trial required the subject to perform 100 button presses, using the non-dominant little finger within 21 seconds, while successful completion of easy-trial required the subject to make 30 button presses, using the dominant index finger within 7 seconds. In EEfRT, subject performance reflects individual differences in the willingness to expend effort for a given level of expected reward utility (reward amount weighted against the probability of winning). Thus, the behavioural aspect of the concept of “wanting” is being manipulated. Indeed, it was found that individuals who reported higher levels of anhedonia exhibited a reduced willingness to make choices requiring greater effort in exchange for greater reward^4^.

#### Real-time fMRI Neurofeedback

Before entering the MR scanner, subjects were reminded of the NF task instructions, and provided with a list including at least three strategies they wish to begin with. The strategies were incorporated into the choice screen by the experimenter using in-house software, that in addition accompanied the preparation of all structural, functional and ROI files needed for the NF training session. Subjects were instructed to move as little as possible.

#### Reward-ML network functional localization

Prior to NF training, subjects performed the MID task, which was employed as a functional localizer for reward-ML NF subjects. Following MID performance, fMRI images were pre-processed using SPM12 (www.fil.ion.ucl.ac.uk/spm). Preprocessing steps included realignment, reslicing and spatial smoothing with an isotropic Gaussian kernel of 4-mm full width at half maximum (FWHM). For each run, a GLM was constructed with anticipation (high, low, neutral), cue response, and consumption (high, low, neutral) as explanatory regressors. Error trials were included as regressors of no interest, along with six head motion parameters. A second (subject)-level GLM was averaged over two runs, from which individual statistical t-maps of reward anticipation contrast (high>neutral reward anticipation) were extracted.

For the bilateral Nac, predefined meta-analytic 8-mm masks (**Figure S1**) were transformed into the subject’s functional native space, by subjecting the ROIs with the inverse transformation of a segmented anatomical image of the respective participant using SPM12 (http://www.fil.ion.ucl.ac.uk) and custom-made Matlab scripts. Within these ROIs, the peak activation of reward anticipation contrast t-maps was identified. Then, 5 mm spheres were constructed around the peak activation voxels, comprising the subject’s individual Nac ROIs.

Due to the VTA’s small size, anatomical complexity and its vicinity to CSF segments, we applied the following iterative algorithm:

First, we multiplied the subject’s MID reward anticipation t-map with the probabilistic VTA map (transformed into the subject’s native space), which was previously used for VTA localization in NF contexts^9^ (see **Figure S1**). The resulting map captured for each voxel the combined likelihood of being related to reward anticipation for the specific participant, and of being located inside the dopaminergic VTA (as opposed to the substantia nigra).

Then, beginning with radius r=3mm (with a 2mm incremental increase for each iteration), we:

a. Constructed a spherical ROI around the peak voxel in the multiplied map, with current radius = r.
b. Masked the spherical ROI from (a) with a binarized version of the probabilistic VTA mask (thus excluding voxels with zero likelihood of being inside the VTA).
c. Checked whether the number of voxels of ROI (b) is less than or equal to 5 voxels. If the number of voxels was lower, (a) to (c) was repeated until the number of voxels in ROI was larger than 5.

This heuristic method, which intersects the likelihood maps of reward anticipation and neuroanatomical probability of VTA localization, with a minimum voxel count of 5, resulted in individual ROIs with voxel count roughly equal to a spherical ROI with a 2.5-3mm radius, with all voxels within the VTA.

Finally, to improve signal-to-noise ratios, an individual CSF probability map was binarized based on a threshold of 0.8 and used to exclude high-probability CSF voxels from all ROIs.

#### Functional localization of randomized networks

To determine whether post-NF immunological effects derive specifically from reward mesolimbic system modulation during NF training, rather than from mere NF practice (which, as mentioned earlier, includes some aspects of reward as well), effects of reward-ML system NF were compared to a control group. Building on our previous theoretical analyses of NF control conditions^10^, we employed a new control condition to determine specificity of NF outcome effects. This control condition was termed Randomized Network NF (**Figure S2**). Participants in this group were randomly allocated to upregulate one of four non-mesolimbic functional networks, each consisting of three 5-mm radius ROIs taken from meta-analyses. ROIs within each network were created as follows: First, the subgroup-specific ROIs of each participant were transformed from MNI to functional native space as described above for the Nac. Then, similarly to reward ML, NF participants, CSF probability maps were binarized based on a 0.8 threshold, and were used to exclude high-probability CSF voxels from the ROIs.

#### NF double-blinding and randomization

Participants, experimenters and outcome assessors were all blinded to NF group allocation throughout the experiment. Specifically, the primary hypothesis of the study (the link between reward-related processes and immune functions) was withheld from participants. In addition, instructions before training did not mention the reinforced neural target or its functional definition, nor did we mention preferred process-specific mental strategies. To ensure experimenter blinding to NF group allocation, the real-time NF software (OpenNFT) was adapted to not present ROIs on the experimenter screen. ROI creation and integration into the NF software were performed automatically in Matlab in a completely blinded fashion, which did not reveal group allocation to experimenters. Participants were randomly assigned to one of the three study groups via in-house Matlab (https://www.mathworks.com) function. These modifications rendered the NF intervention indistinguishable across groups. Outcome assessors of neural, mental strategies and immunological outcome measures were blinded throughout data analysis (group allocation was coded as either A,B or C, and each group’s identity (reward-ML-NF, Randomized-network-NF and no-NF) was revealed only after all outcome measures and effects were assessed.

#### Real-time fMRI neurofeedback set-up and preprocessing

A PC running "Open NFT״ software^11^ (http://opennft.org/) received each functional volume from the MR console computer as raw .dicom file, and performed the standard OpenNFT preprocessing steps on the data, based on SPM12 functions (www.fil.ion.ucl.ac.uk/spm) that were adapted for real-time purposes. Specifically, spatial realignment to the first image, estimation of six movement parameters (translation and rotation), reslicing, and spatial smoothing with an isotropic Gaussian kernel with 4-mm full width at half maximum (FWHM) were performed. First order auto-regressive correction was applied to reduce temporal autocorrelation caused by physiological noise^12^, and an incremental general linear model (iGLM) was used to remove residual motion and linear trends^13^. Spike detection and high-frequency noise removal were performed through a modified Kalman filter^14^. Whole-brain activation maps were estimated using iGLM statistical analysis^15^, with NF task protocol conditions (**Figure 1b**) as regressors of interest. ROI BOLD levels were calculated for each ‘Rest’ and ‘Upregulate’ period at the termination of the regulation screen, and transformed in real-time to feedback scores, as detailed below. Full details of OpenNFT processing steps can be found in ref^11^.

#### On-line calculation and presentation of feedback

The intermittent feedback was calculated for each cycle as follows: First, to account for the Hemodynamic Response Function (HRF) delay, the first four EPI functional volumes of Rest and upregulate epochs were discarded from analysis (TR=2 s.). Then, for each ROI, median level of each epoch was calculated and scaled according to the following formulas:

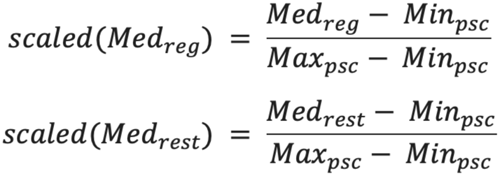

With and being the minimum and maximum BOLD% values registered up until the current TR. Then, the regulation effect for the current trial was calculated as the difference between scaled values of ‘Upregulate’ and ‘Rest’ epochs:

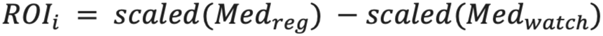

This results in a number between -1 and 1, indicating the scaled difference between regulation and rest screens, with 1 indicating a maximum upregulation effect, and -1 a maximum downregulation effect based on non-parametric statistics. Following this, the regulation effects of the entire target network were calculated as the mean regulation level across the three network ROIs:

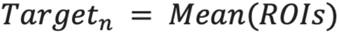

These regulation effects were transformed into the feedback level, defined as a number between 1 and 10, by multiplying by 10, and imposing a lower limit of 1. Accordingly, all downregulation effects (and very slight up-regulation effects proportional to 0-0.1 regulation levels) were similarly depicted as 1. Finally, the feedback score was displayed to the participant next to an image of a smiley face, while the strength of the smile was proportional to the feedback level (1 – a neutral smiley; 10 – a maximally smiling face).

#### Documentation of online mental strategies and updating of choice screens (Figure S3)

Before the first run of a NF training session, participants defined and named three strategies they planned to employ during the next NF run based on their previous experience. The experimenter incorporated the new strategy names into a run-specific choice screen via in-house scripts that were uploaded by the OpenNFT real-time software as the screen image for choice epochs. Critically, an additional open category (“other”) always appeared as a fourth option. By including such an option, we ensured that participants were not restricted to their predefined options, allowing them to act on any momentary intuitions during runs. At the beginning of each trial, participants chose via key press which strategy they wished to employ in the next regulation screen. Between runs, subjects were given the opportunity to replace one or more of the predefined strategies, or to remain with the same three options, according to their accumulated experience (see “between-run instructions” in ‘Supplementary Materials’ section). At the end of the training session, an output log file was exported, containing the names of each strategy employed, during each trial of the training session.

#### Characterization of mental strategies

To characterize mental strategies logged during choice screens for each trial, we developed a new classification method aimed at exhaustively assessing the mental features of strategies, while placing special focus on reward-related and affective features that could arise during reward-ML NF regulation periods.

The Mental Strategies Questionnaire for NeuroFeedback (MSQ-NF) is constructed hierarchically with general meta-categories each containing several features, resulting in a total of 45 features (**Figure S3**). Features were selected to account for the main mental modalities, contents and dimensions as defined in the affective and cognitive psychological literature, such as exteroception and interception, affect (e.g. discrete emotions, arousal and valence dimensions), imagery and memory features, as well as lingual, motor, arithmetic and social aspects.

The documentation procedure described earlier produced a trial-locked list of applied strategies named by each subject, which appeared on their choice screens. Using this list as a mnemonic aid of trial-specific choices, at the end of each NF training session participants: (i) provided a detailed verbal description of each strategy headline from the documentation list, and (ii) filled out the MSQ-NF for each applied strategy via ‘Qualtrics’, by stating whether the strategy was or was not characterized by each MSQ feature. This labelling process results in a binary matrix, with NF trials in rows and MSQ features in columns. The trials-by-MSQ features matrix was used to characterize the changing mental features across all Upregulate’ epochs of the practice.

#### Questionnaires

To further assess the involvement of trait motivational capacities and tendencies in the physiological regulation of immune functions, participants filled out three trait questionnaires:

- Sensitivity to Punishment and Sensitivity to Reward Questionnaire (SPSRQ)^3^, measuring the behavioural appetitive system and reward sensitivity. SPSRQ is a 24-item questionnaire with two subscales – reward and punishment. It is one of the most widely used measures to assess individual differences in motivational tendencies, including behavioural inhibition in response to novelty or punishment cues and cognitive worry in response to failure or punishment cues. Response to each item is provided in a yes/no dichotomous manner.
- ‘Neuroticism-Extraversion-Openness Five-Factor-Inventory (NEO-FFI)^2^ measuring five factors of personality traits, which includes subscales relating to reward and motivational behaviour. NEO-FFI is a widely used self-reported psychological personality inventory, which includes 60 questions (an abbreviated version of the 240-item NEO-PI-R). This ‘Big Five’ model includes the traits: extraversion, neuroticism, openness, agreeableness and conscientiousness, and comprises a meeting point for disparate human personality models. Its scales have been associated with various physiological and psychiatric features as well as with motivational tendencies^16^.
- the Tridimensional Personality Questionnaire (TPQ)^1^, a personality inventory measuring three major personality dimensions: Novelty Seeking (NS), Harm Avoidance (HA) and Reward Dependence (RD).

These three questionnaires were previously used by Gonen and colleagues^17^ to classify individual participants via K-means clustering procedure as having avoidance or approach tendencies. The three questionnaires were filled at the beginning of the first experimental session.

#### fMRI acquisition

All scans were performed on a 3.0 Tesla Siemens MRI system (MAGNETOM Prisma, Germany) using a 64-channel head coil. Structural scans included a T1-weighted 3D Sagittal MPRAGE pulse sequence (TR/TE = 1980/2.62 mesc., flip angle = 8°, pixel size = 0.9X0.9mm, FOV = 224×224 mm, slice thickness = 0.9 mm).

Functional whole-brain scans were performed in interleaved order with a T2*-weighted gradient multi-echo planar imaging pulse sequence (TR/TE=2000/35ms, IPAT acceleration factor=2; flip angle=84°, FOV=220×220mm, matrix size = 96*96, voxel size: 2X2 mm, slice thickness = 2mm, no gap, 56 slices per volume).

#### fMRI pre-processing

Neuroimaging data were converted to BIDS format and pre-processed via fMRIprep software package (version 20.0.2)^18^, and then pre-processed as described below:

#### fMRI anatomical preprocessing

Anatomical images were corrected for intensity non-uniformity (INU) with ‘N4BiasFieldCorrection’, distributed with ANTs 2.2.0. The T1w-reference was then skull-stripped with a Nipype implementation of the ‘antsBrainExtraction.sh’ workflow (from ANTs), using OASIS30ANTs as target template. Brain tissue segmentation of cerebrospinal fluid (CSF), white-matter (WM) and grey matter (GM) was performed on the brain-extracted T1w using ‘fast’ (FSL 5.0.9, fsl_fast). A T1w-reference map was computed after registration of T1w image (after INU-correction) using ‘mri_robust_templatè (FreeSurfer 6.0.1, fs_template). Volume-based spatial normalization to one standard space (MNI152NLin2009cAsym) was performed through nonlinear registration with ‘antsRegistration’ (ANTs 2.2.0), using brain-extracted versions of both T1w reference and the T1w template. The following template was selected for spatial normalization: ICBM 152 Nonlinear Asymmetrical template version 2009c; Template used for anatomical alignment and registration: MNI152NLin2009cAsym.

#### fMRI functional preprocessing

The following preprocessing was performed: First, a reference volume and its skull-stripped version were generated using a custom methodology of fMRIPrep. A B0 nonuniformity map (or fieldmap) was estimated based on a phase-difference map calculated with a dual-echo GRE (gradient-recall echo) sequence, processed with a custom workflow of SDCFlows inspired by the ‘epidewarp.fsl’ script (http://www.nmr.mgh.harvard.edu/~greve/fbirn/b0/epidewarp.fsl) and further improvements in HCP Pipelines. The fieldmap was then co-registered to the target EPI (echo-planar imaging) reference run and converted to a displacement field map (amenable to registration tools such as ANTs) with FSL’s ‘fuguè and other ‘SDCflows’ tools. Based on the estimated susceptibility distortion, a corrected EPI (echo-planar imaging) reference was calculated for a more accurate co-registration with the anatomical reference. The BOLD reference was then co-registered to the T1w reference using ‘flirt’ (FSL 5.0.9) with the boundary-based registration (bbr) cost-function. Co-registration was configured with nine degrees of freedom to account for distortions remaining in the BOLD reference. Head-motion parameters with respect to the BOLD reference (transformation matrices, and six corresponding rotation and translation parameters) were estimated before any spatiotemporal filtering using ‘mcflirt’ (FSL 5.0.9). The BOLD time-series (including slice-timing correction when applied) were resampled onto their original, native space by applying a single, composite transform to correct for head-motion and susceptibility distortions. These resampled BOLD time-series will be referred to as ‘pre-processed BOLD in original space’, or simply pre-processed BOLD. The BOLD time-series was resampled into standard space, generating a pre-processed BOLD run in MNI152NLin2009cAsym space. First, a reference volume and its skull-stripped version were generated using a custom methodology of fMRIPrep. Several confounding time-series were calculated based on the pre-processed BOLD: framewise displacement (FD), DVARS and three region-wise global signals. FD and DVARS were calculated for each functional run, both using their implementations in Nipype (following the definitions by power_fd_dvars). The three global signals were extracted within the CSF, the WM, and the whole-brain masks. Additionally, a set of physiological regressors was extracted to allow for component-based noise correction (CompCor). Principal components were estimated after high-pass filtering the pre-processed BOLD time-series (using a discrete cosine filter with 128s cut-off) for the two CompCor variants: temporal (tCompCor) and anatomical (aCompCor). tCompCor components were then calculated from the top 5% variable voxels within a mask covering the subcortical regions. This subcortical mask was obtained by heavily eroding the brain mask, which ensures it does not include cortical GM regions. For aCompCor, components are calculated within the intersection of the aforementioned mask and the union of CSF and WM masks calculated in T1w space, after their projection to the native space of each functional run (using the inverse BOLD-to-T1w transformation). Components are also calculated separately within the WM and CSF masks. For each CompCor decomposition, the k components with the largest singular values are retained, such that the retained components’ time series are sufficient to explain 50 percent of variance across the nuisance mask (CSF, WM, combined, or temporal). The remaining components are removed from consideration. The head-motion estimates calculated in the correction step were also placed within the corresponding confounds file. The confound time series derived from head motion estimates and global signals were expanded with the inclusion of temporal derivatives and quadratic terms for each (confounds_satterthwaite_2013). Frames that exceeded a threshold of 0.5 mm FD or 1.5 standardised DVARS were annotated as motion outliers. All resamplings could be performed with a single interpolation step by composing all the pertinent transformations (i.e. head-motion transform matrices, susceptibility distortion correction when available, and co registrations to anatomical and output spaces). Gridded (volumetric) resamplings were performed using ‘antsApplyTransforms’ (ANTs), configured with Lanczos interpolation to minimize the smoothing effects of other kernels. Non-gridded (surface) resamplings were performed using ‘mri_vol2surf’ (FreeSurfer).

Finally, fMRIprep pre-processed images were spatially smoothed using a Gaussian kernel at 4 mm FWHM, via nilearn package (Abraham et al., 2014) in Python.

#### Immunological assessments

Blood samples were collected from all participants at the TLVMC (Ichilov). Blood withdrawal time points (TP) were scheduled before the 1st and 4th NF sessions (TP1, TP4), and 14 and 28 days following HBV vaccination (TP5, TP6, respectively) for the measurement of HBV antibody levels. A subset of subjects (N=34) arrived for TP-7 to provide a blood sample 3 months following HBV vaccination, to examine long-term immunological effects. Following blood withdrawal, samples were centrifuged and plasma was extracted, aliquoted, and then stored at -80°C. Blood samples were transferred to the Rolls lab for antibody titer analysis, as described below.

#### Outcome measures

The primary neural outcome measure of the study was reward-ML activations during the NF task (described in section “ROI analysis: characterization of regulation effects”). There were two main immunological outcome measures: post-vaccination HBVab change (described in “Immunological analysis: HBV antibody change”) and inflammatory activity manifested in changes in blood cytokines levels. The latter is outside the scope of the current paper, which focuses on vaccination efficiency. There are two types of secondary outcome measures for the study. The first is a long-term assessment of vaccination efficiency measured as HBVab levels three months following HBV vaccination. The second type consists of variables that could explain the main outcomes of the intervention (i.e. the primary neural and immunological outcome measure). These were: reward-related mental features of mental strategy application during NF regulation; neurobehavioural indices of trait reward anticipation and consumption measured by the MID task before NF training; trait behavioural incentive motivation as measures by the EEfRT before NF training; and motivational tendencies of participants as measures by K-means cluster analysis of designated questionnaires data. The statistical assessment of all outcome measures is specified below.

#### fMRI-NF ROI analysis: characterization of regulation effects

Neuroimaging data were analysed via the FMRIB software library (FSL)^19^, as specified below.

##### Functional localization of reward-ML ROIs

To define the VTA and Nac ROIs, we analyzed our functional localizer – the MID task. First, to validate the online functional localization procedure described earlier, we examined whether the High>Neutral reward anticipation contrast, used online to functionally localize the reward ml system at the individual subject level, indeed elicits significant activations in mesolimbic regions on the group level. Based on group-level analyses, we extracted ROIs of the reward-ML network for the characterization of neural effects during NF training. While the offline subject-level analyses conducted offline were essentially equivalent to the real-time functional localization method, they benefitted from superior offline preprocessing via fMRIprep (described above), and from the statistical power the group analysis yields, resulting in a more robust localization of reward-ML ROIs that are consistent across subjects.

**Figure S1** depicts cluster-corrected (p(FWE)<0.05) whole-brain BOLD activations during high vs neutral reward anticipation. As reported in previous studies^7,20^, reward anticipation yielded significant activations in mesolimbic and cortical regions, including the VTA and the bilateral Nac, the Anterior Cingulate Cortex (ACC), and the thalamus.

As for the online functional localization procedure, offline reward-ML ROIs were created by intersecting the predefined meta-analytic masks (**Figure S1,** overlaid in white) with the BOLD activity patterns in the High>Neutral reward anticipation contrast (Orange-yellow colour dimension), resulting in a reward anticipation-related mesolimbic ROIs of the VTA and bilateral Nac, presented in Figure S1, lower panel (See figure legend for a description of intersection steps). These ROIs were used in the fMRI-NF analysis described below.

##### Production of whole-brain activity maps

First-level data processing was carried out using FEAT (FMRI Expert Analysis Tool) Version 6.00, part of FSL (www.fmrib.ox.ac.uk/fsl). Time-series statistical analysis was carried out using ‘FILM’ with local autocorrelation correction^21^. The general linear model of a NF run included regressors of interest of the four task conditions: (1) Choice (8 s), (2) Rest (20 s), (3) Upregulate (40 s), and (4) feedback (8 s). To account for noise, confound regressors from the fMRIprep preprocessing were included as covariates of no interest. The following nuisance regressors were included: six head movement regressors (translation and rotation), framewise displacement predictor, first six anatomical CompCors, as well as mean CSF and white matter global signal predictors. Additionally, we introduced scrubbing regressors into the noise model based on framewise displacement variable, regressing out time points with FD > 0.5. Runs containing more than 20% scrubbed timepoints, were completely excluded from further analyses due to high levels of head movements. For NF training data, no runs exceeded this threshold. All predictors were convolved with a canonical hemodynamic response function, and data were subjected to FSL default high-pass filter cutoff (128 s).

The three main Contrasts of Parameter Estimates (copes) were then computed: “Choice”, which represented activity patterns during choice of mental strategies; “Regulate>Rest”, which represented regulation effects – the difference in activity patterns between Upregulate and Rest screens; and “Feedback”, which represented activity pattern during rewarding feedback regarding regulation success during feedback screens.

Effects of each session were averaged using a fixed effects model, by forcing the random effects variance to zero in FLAME (FMRIB’s Local Analysis of Mixed Effects)^21–23^. Finally, group-level effects for session 4 were carried out using FEAT: Z (Gaussianised T/F) statistic images were thresholded using clusters determined by Z>3.1 and a (corrected) cluster significance threshold of P=0.05.

##### NF reward-ML network regulation effects - group differences

To assess NF regulation effects across groups and sessions, a linear mixed effects model was conducted using the ‘lmer’ function from the lme4 package in R version 4.2.1 (https://www.R-project.org/)^24^. Models were used to estimate differences in activity in the VTA and the Nac ROIs across groups and sessions. Importantly, since one third of subjects began NF training only in session 2, session 1 was discarded from the analysis of group differences (as including it prevented the model from converging due to failure of assessing random effects).

##### Quantification of individual regulation effects for brain-immune correlation analyses

Our pre-registered brain-immune correlation hypothesis describes the neural measure as ROI activity levels in session-4, without mentioning if and how baseline regulation levels (i.e. at the beginning of the training) will be taken into account. We note that, similar to the characterization of the immunological effect as HBVab *change* between post-vaccination and baseline HBVab levels, we applied a common individualized measure of subject-specific regulation effects, by quantifying the slope (increase or decrease) of ROI activity across all runs, from session 1 to session 4^27–30^. Accordingly, regulation effects per participant were calculated as the slope of a linear regression of ‘Upregulate>Rest’ BOLD percent signal changes in each ROI across runs, resulting in two regulation effect measures per individual, one for the VTA and one for the Nac. For non-mesolimbic control regions, ‘Upregulate>Rest’ effects in ROIs of each functional network in the randomized-network control group were calculated as in the reward-ML ROIs and averaged within each run to constitute one mean regulation value of the whole network. Then, individual regulation effects were calculated as described for the reward-ML ROIs.

#### Immunological analysis: HBV antibody change

Antibody titer to Hepatitis B surface antigen (anti-HBs) were determined, using a Chemiluminescent Microparticle Immunoassay (CMIA) located in the Virology lab at the Rambam Health Care Campus. In any case antibody titer exceeded the maximal detection range (1000 mU/ml), the sample was diluted in antibody-negative plasma (at a ratio of 1:10-1000). The samples were also quantitatively tested for hepatitis B surface antigen (HBsAg), to exclude any participant who might have been infected with Hepatitis B at the time of vaccination. No participant was excluded based on this criterion.

HBVab levels were log-transformed. Then, post-vaccination immune response was represented as the difference between mean HBVab levels during sessions 5 and 6 and mean baseline measurements, at sessions 1 and 4. This resulted in a single value per participant that quantified immune dynamics following NF induction and HBV vaccination coupling. Twelve subjects (evenly dispersed between all groups) did not respond to HBV vaccination (according to the standard clinical immunization threshold of HBVab levels below 10 mU/ml^25^) and were therefore excluded from further analyses. Of note, this data exclusion criterion was no pre-registered. Nonetheless, as the lack of an immunological response to HBV vaccination is a phenomenon that was previously documented, we assumed our vaccination-induced manipulation failed for these subjects regardless of NF training (indeed, 5 out of 12 non-responders were from the no-NF group). Therefore, non-responders were excluded from any immunological or brain-immune association analyses.

To test differences between groups, we conducted a one-way ANOVA, using f_oneway function from the Scipy package^26^ in Python (http://www.python.org) version 3.8.12, with group as between-subjects factor (reward ml NF/rand. ROI NF/no NF) and post-vaccination HBVab log change as dependent variable. Correlations between individual ROI regulation effects and HBVab change were determined using Python version 3.8.12.

#### Neurobehavioural measures of reward anticipation and consumption during MID task

##### Extraction of Whole-brain BOLD activation maps

For each MID run (out of two experimental runs), first-level GLM for an event-related design was carried out. Regressors of interest included the three anticipation phases: anticipation of no reward (0 NIS), anticipation of low reward (2 NIS) and anticipation of high reward (10 NIS). Similarly, for outcome phases, we included one regressor per condition (high, low and neutral outcome regressors). Additionally, for the low and high reward conditions, the two outcome regressors were parametrically modulated by the actual outcome amount received at each trial. Target presentation (one regressor) and anticipation, and target and outcome phase during error trials (3 regressors) were modeled as regressors of no interest. In total, the first-level model included 12 regressors of interest. A canonical HRF was applied for convolving explanatory variables. To account for noise, confound regressors from the preprocessing step were included as covariates of no interest. The noise model was identical to the NF task analyses described earlier. One subject was excluded from analyses due to extreme head movement (i.e. > 20% scrubbed time points according to a cutoff of FD>0.5). Three additional subjects did not complete two full experimental runs and were therefore excluded as well, resulting in 81 subjects for group analyses.

COPEs (Contrasts of Parameter Estimates) for reward anticipation and reward outcome were calculated. For reward anticipation, we calculated the contrast anticipation of high reward versus anticipation of no reward, which was used for the functional localization process of the reward-ML network described earlier. For the reward outcome, we included in the first-level model the parametric modulator for high reward (in addition to the general reward consumption regressors). Then, effects per participant were averaged across the two runs using second-level analysis. This was carried out using a fixed effects model, by forcing the random effects variance to zero in FLAME.

Third-level effects were calculated to assess reward anticipation and outcome group effects. For each contrast, individual copes of all participants were included in a mixed-effects model using FSL FEAT Version 6.00. Z (Gaussianised T/F) statistic images were thresholded using clusters determined by Z>3.1 and a (FWE corrected) cluster significance threshold of P=0.05.

##### ROI analyses

Neural reward responsivity measures were extracted for reward anticipation and reward consumption from first-level whole-brain contrast maps via FSL “featquery” and converted to percent signal change. For reward anticipation, activations were extracted from the VTA and Nac. For reward outcome, a ventromedial prefrontal cortex (vmpfc) ROI was used. Vmpfc was defined as a 5mm radius spherical ROI around peak activation in the medial pfc section in the whole-brain reward outcome group effects (MNI: X=-4, Y=59, Z=-1). Associations between these two neural measures and immune effects were estimated via correlational analyses reported in the main text and in ‘Supplementary Information’ section

#### Behavioural incentive motivation during EEfRT

In line with previous studies that employed EEfRT^4,20^, mean proportions of hard-task choices were created for all subjects across each level of probability. Proportions of hard-task choices were averaged over all reward probabilities per individual as a measure of effort expenditure. This measure was used for correlational analyses reported in the ‘Supplementary Information’ section.

#### Mental Strategies characterization

Choice screen log files of choices and trial-locked MSQ labelling were integrated across all sessions and participants, resulting in 3240 NF trials across 45 MSQ features (between 45 and 60 trials per participant). A value of ‘1’ indicated that a particular feature characterized a trial, while ‘0’ indicated that it did not, and ‘Nan’ values indicated neither, either based on the participant’s explicit judgment or if labelling was not performed by the participant. To answer our research questions that focused on regulation prior to HBV-vaccination, only MSQ data from the 4th session were analyzed, which included 962 NF trials. Features that characterized less than 10% of trials were considered irrelevant to our NF training setting, and thus were discarded from further analyses, resulting in 29 MSQ features. Excluded features were: ‘visceral sensations’, ‘low arousal’, ‘other bodily sensation’, ‘tactile exteroception’, ‘pulse’, ‘breathing’, ‘taste imagery’, ‘other emotion’, ‘muscle sensation’, ‘anger’, ‘taste exteroception’, ‘disgust’, ‘vision exteroception’, ‘fear’, ‘auditory exteroception’ and ‘smell exteroception’. These data were used to assess the link between reward-related mental features and reward-ML activity as described in the ‘Results’ section.

#### K-means clustering of motivational tendencies from questionnaires data

To classify motivational tendencies, participants were divided into two groups characterized by either approach or avoidance tendencies, based on a two-step K-means clustering of questionnaire data. 70 subjects were included in the analysis (15 subjects did not complete at least one of the three questionnaires included in the analysis, and were therefore excluded). A two-step non-hierarchical K-means cluster analysis was performed via Python’s ‘*KMeans’* function from *sklearn* package^31^, imposing two distinct clusters based on the clustering used in^17^. First, the analysis was performed based on normalized scores of all traits measured by thirteen sub-scales derived from three personality questionnaires (NEO-FFI, SPSRQ and TPQ) per participant. The traits that did not demonstrate a significant difference between the two clusters based on an independent t-test conducted as part of the two-step K-means procedure were excluded. Excluded subscales were openness from the NEO-FFI, Novelty Seeking from the TPQ, and Reward Sensitivity from the SPSRQ. Next, a second K-means cluster analysis was performed (again, with two clusters) which included the subscales in which a significant difference was found between clusters: Neuroticism, Extraversion, Conscientiousness and Agreeableness from NEO-FFI, Reward Dependence and Harm Avoidance from TPQ, and Punishment Sensitivity from SPSRQ. This process resulted in two clusters characterized by distinct tendencies of approach (N=43) or avoidance (N=27). Based on this clustering, we assessed the link between approach/avoidance motivational tendencies, reward-ML modulation effects and adaptive immune functions.

## Data availability statement

Requests for de-identified data can be directed at the corresponding author (Talma Hendler). All requests for data sharing will be reviewed by the Tel-Aviv Sourasky Medical Center Institutional Review Board (IRB) committee, to verify whether the request is subject to any intellectual property or confidentiality obligations. Requests will be reviewed on the basis of scientific merit, ethical review, available resources and regulatory requirements, and will be responded within 90 days. After approval of a proposal, anonymized individual-level data will be made available for reuse in accordance with the signed consent IRB form. A signed data access agreement with the collaborator is required before accessing shared data.

## Notes

### Clinical Trial

NCT03951870

### Funding Statement

This study was funded by Joy Ventures foundation and by KAMIN program in the Israeli
ministry of innovation. N.L. would like to thank the Teva Bio-Innovation Forum.

### Author Declarations

the IRB of the Tel-Aviv Sourasky Medical Center gave ethical approval for this work.

