## Supplementary Information for "Upregulation of Reward Mesolimbic Activity via fMRI-Neurofeedback Improves Vaccination Efficiency in Humans"

Figure S1: Functional localization of the reward mesolimbic network

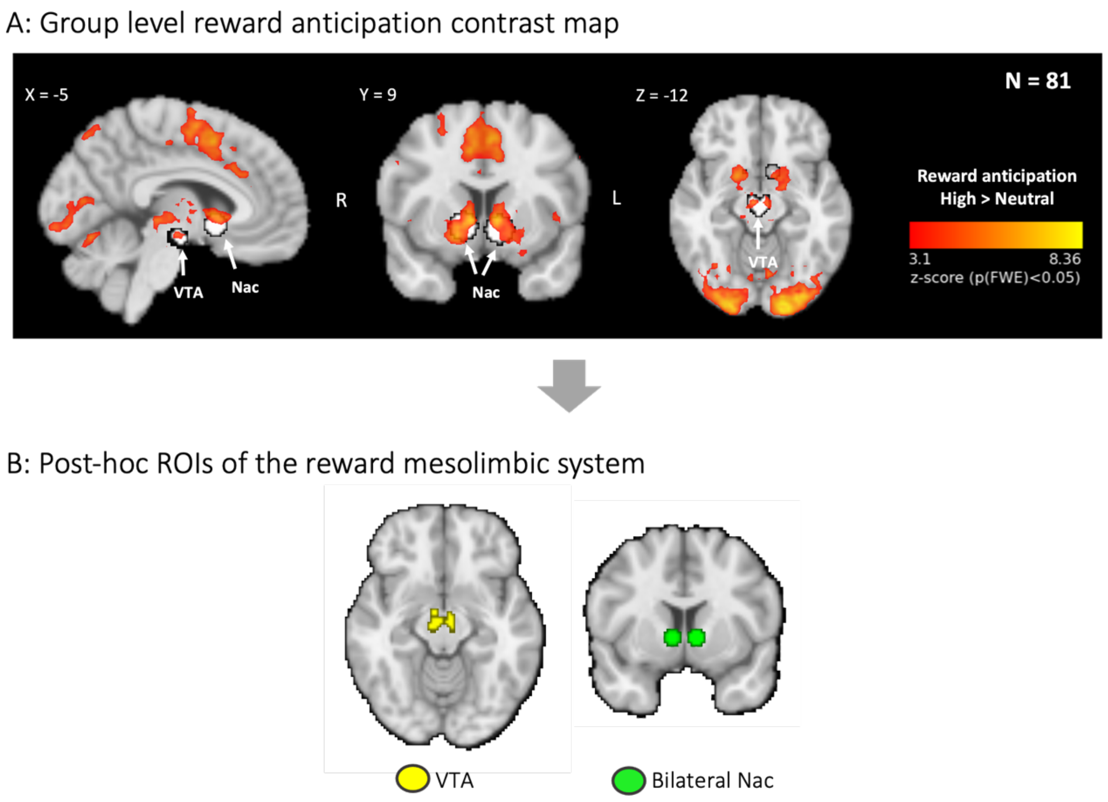


**Figure S1: Offline functional localization of the reward mesolimbic network. A.** Whole-brain results for the high vs neutral reward anticipation contrast. The overlaid predefined meta-analytic masks of the mesolimbic system, taken from refs[1,2], are shown in white. Abbreviations: Nucleus Accumbens (Nac); Ventral Tegmental Area (VTA). Statistical threshold was set at a cluster-level p(FWE)<.05. **B.** the offline functional localization of the reward mesolimbic regions. For the VTA, the predefined anatomical mask[2] was intersected with the high>neutral reward anticipation maps presented in A. For the bilateral Nac, a 5 mm sphere was created around the peak activation voxel within the 8mm predefined Nac masks (defined based on reward anticipation meta-analysis[1]).

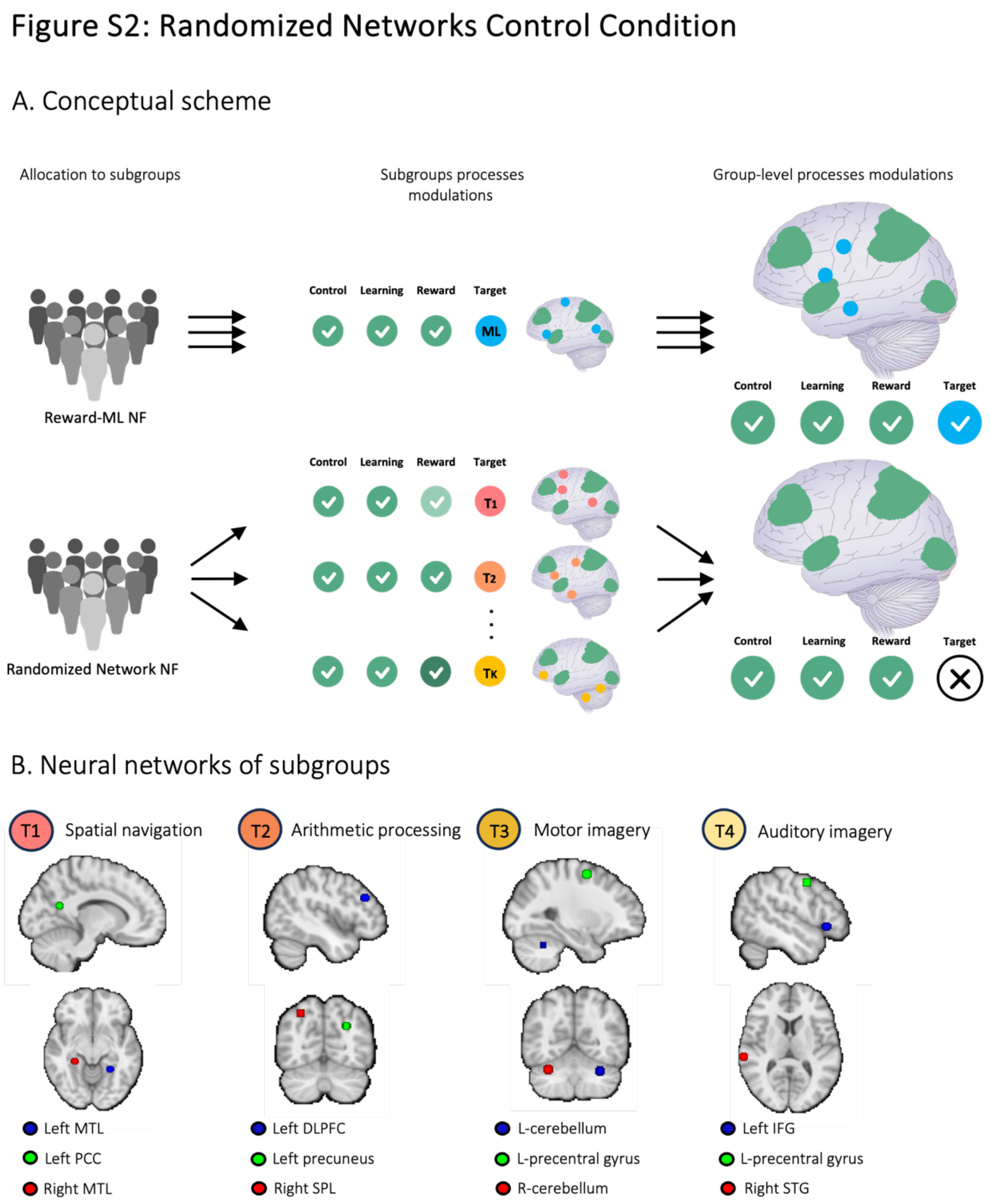


**Figure S2: Randomized Network control condition**. **A.** Conceptual scheme: In randomized ROI-NF, each participant is randomly assigned to one of K sub-groups of different functionally defined neural targets (colored in pink, orange, and yellow). While in each sub-group, a specific neural target is modulated along with the general task processes (colored in green – control, learning and reward processes), group-level modulations (right panel) include only the averaged (non-specific) general processes common to all subgroups (i.e. the target process specific to each subgroup averaged over the whole group). Conversely, in a typical experimental NF group, all participants modulate both the general and target processes (coloured in green and blue, respectively). Therefore, group-level modulations include both the general processes and the reoccurring target process modulations. Consequently, differences in outcome effects (such as immune function) between groups can be attributed solely to the target-specific effects (described in ref[3]). **B.** targets of the randomized ROI-NF control condition. All target networks were selected from meta-analyses of fMRI studies on specific functional processes. MNI coordinates: T1. Spatial navigation network[4]: right medial temporal lobe: 26, -35, -11; left medial temporal lobe: -26, -47, -9; left posterior cingulate: -15, -59, 19; T2. Arithmetic processing network[5]: right SPL: 29, -66, 49; left dlpfc: -45, 32, 29; left precuneus: -28, -71, 33; T3; Motor imagery network[6]: right cerebellum:

32, -62, -28; left cerebellum: -32, -56, -30; left precentral gyrus: -26, -2, 58; T4. Auditory imagery network[7]: right STG:64, -30, 9; left IFG: -48, 24, -5; left precentral gyrus: -52 1 47.

#### Figure S3: Documentation and Characterization of Mental Strategies

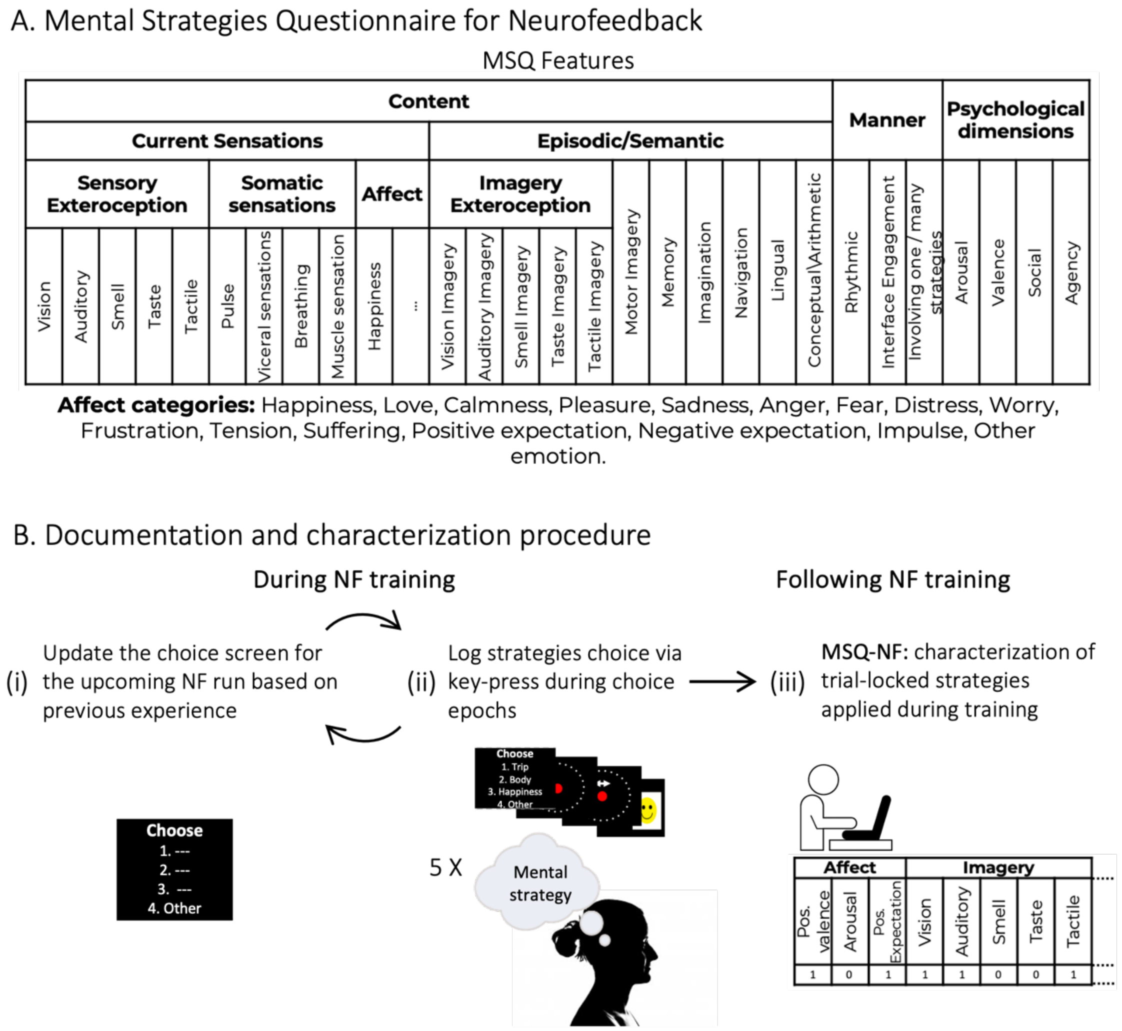


**Documentation and characterization of mental strategies. A**. Before each NF run (comprised of five consecutive trials), the subject predefines three strategies they may wish to employ on the following run regulation screens, based on their previous experience (with a fourth "other" set option that enables choosing strategies conceived on-the-fly based on real-time intuitions). The experimenter updates the screen with the strategy names provided by the subject. **B.** During task choice epochs, the subject indicates which strategy they wish to employ in the next regulation screen via key-press, with three predefined options and a fourth open category ("other"). Thus, choices of strategies are logged for each trial. This process repeats itself for as many runs as needed per session. **C.** Following practice, participants fill out the MSQ-NF for each strategy applied during the session, based on their input of strategies used. "Other" strategies as well as those with predefined names are verbally described, and then the strategy is labelled across the MSQ space.

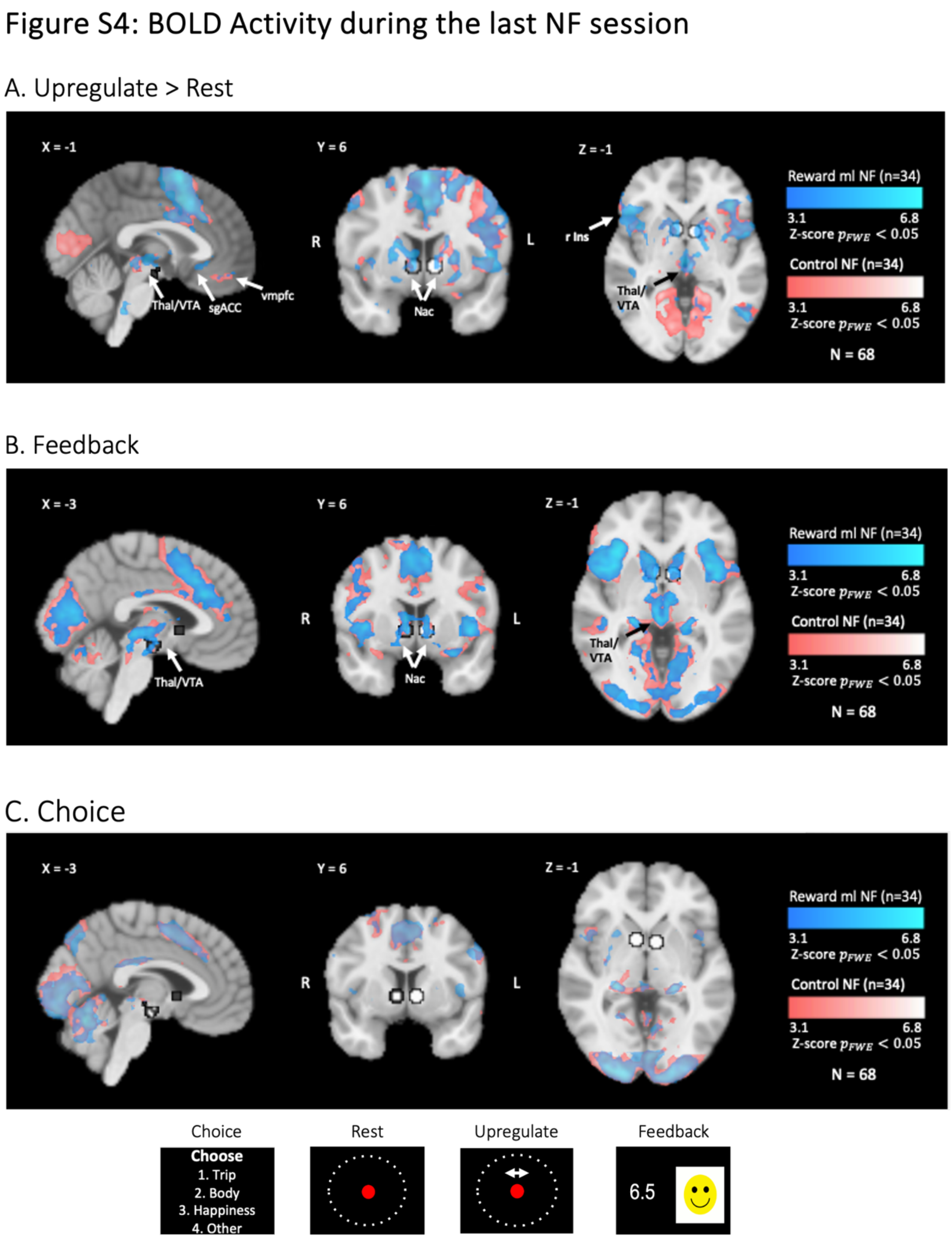


**Figure S4: BOLD activity during the last NF session across conditions and groups. A.** 'Regulate>Watch' contrast, showing differing regulation effects per group (reward ml NF – light blue, randomized ROI NF – pink). **B.** 'Feedback' contrast. **C.** 'Choice' contrast. Abbreviations: right Insula (r Ins); subgenual ACC (sgACC); Thalamus (Thal), ventromedial prefrontal cortex (vmpfc). The VTA and bilateral Nac ROIs are overlaid in white.

#### Supplemental Results

##### Links between mental features and ROI modulation levels

To further validate the results presented in the main text associating positive expectations with VTA modulation levels, we conducted a logistic regression analysis with positive expectation (logit-transformed) frequency as the dependent measure, and grouping of modulation effects (high vs low) in both mesolimbic targets (VTA and Nac) as predicting factors, as well as a constant intercept term. Results revealed that indeed, only VTA modulation significantly predicted the frequency of positive expectation features (VTA: z=4.29, p<.0001; Nac: z=0. and randomized network modulation grouping conducted only on participants 286, p=.775, N=60). Moreover, to test the efficiency of our approach, we compared VTA from the randomized networks group. This analysis exhibited similar results (albeit weaker, due to lower statistical power), with VTA significantly predicting positive expectation frequency (z=2.43, p<.015, N=30) and a marginally significant effect in the opposite direction for randomized networks (z=-1,86, p=.062). We conducted a similar logistic regression analysis, but with trial-specific ROI activity instead of categorical (high vs low) activity levels based on a general group median split, to have a more fine-grained measure of ROI activity per trial. ROI activity levels were extracted from a General Linear Model (GLM) with trial-specific predictors of each condition instead of the standard averaging within-run across conditions. Other analysis steps were similar to ROI analyses described in the 'Online Methods'. However, these analyses did not show significant results

##### Lack of correlations between trait motivational measures and post-vaccination immune response

The VTA-Immune correlation could result from individual differences in motivational traits. Specifically, it could be that regardless of the temporal contingency between fMRI-NF training and the HBV vaccination, participants with strong neural and/or behavioural motivational tendencies would exhibit both higher reward-ML upregulation effects during fMRI-NF training, and an enhanced post-vaccination HBVab change compared with those with weak motivational tendencies.

To test this possibility, we examined whether post-vaccination HBVab change and reward-ML regulation effects were correlated with gold-standard metrics capturing individual variability in motivational traits.

During session 1, participants from all study groups performed the EEfRT before and the MID task during fMRI scanning. In addition, participants filled three questionnaires capturing motivational tendencies (NEO-FFI, SPSRQ, and TPQ). Individual neural measures of reward anticipation were extracted from the MID task from the Nac and the VTA via ROI analyses. For reward consumption, we extracted MID activation from the ventromedial prefrontal cortex (vmpfc) (see 'Online Methods'). In addition to group-level analysis of reward anticipation (reported in **Figure S1**) we conducted a group-level GLM constructing a z-scored map for reward consumption. Analyses revealed a distinct network that has been previously associated with reward consumption during MID task [1,8], including the medial prefrontal cortex, bilateral Nac and ventral pallidum, as well as visual regions. Then, we constructed a spherical ROI with 5 mm radius around the peak activation of the medial pfc cluster (MNI coordinates: X=-4, Y=59, Z=-1), and conducted an ROI analysis to extract individual neural responses to reward consumption.

We calculated Pearson's correlations between neural responses to reward consumption and anticipation, with post-vaccination HBVab change (N=81 from all groups) and with individual VTA upregulation effects (N=67 from NF groups). Analyses did not reveal significant correlations between reward anticipation and consumption and HBVab change (Nac reward anticipation and HBVab change: r=0.13, p=0.268; VTA reward anticipation and HBVab change: r=0.08, p=0.472; vmpfc reward consumption and HBVab change: r = 0.06, p=0.582) nor between VTA reward anticiptation and VTA upregulation effects (r=0.175, p=0.155).

.

Next, we examined whether effort expenditure as measured in EEfRT relates to HBVab change or VTA upregulation effects. The individual probability to select a hard task across varying levels of reward was extracted per subject from EEfRT performance (see 'Online Methods'), and was correlated with regulation effects and peripheral immune functions. Similar to MID measures, we did not find any significant correlations with these neural and immunological markers (Effort expenditure and HBV change: r=-0.05, p=0.661; effort expenditure and VTA upregulation effect: r=-0.05, p=0.680)

Finally, to capture participants' behavioural motivational tendencies, we conducted a 2-step K-means cluster analysis, conducted on 70 subjects with qualified questionnaire data (see 'Online Methods'). This analysis revealed two clusters that significantly differed in various subscales relating to reward and motivation as quantified by TPQ, SPSRQ and NEO-FFI questionnaires, with 43 subjects classified as exhibiting approach tendencies, and 27 subjects classified as exhibiting avoidance tendencies. Following classification, we first sought to examine the convergent validity of the clustering procedure. In a previous work that used a similar approach, clusters were associated with Nac responses to reward anticipation in a motivational task[9]. Similarly, we examined whether clusters differed in Nac response to reward anticipation during MID task. Independent t-test revealed a marginal difference between clusters ($t(66) = 1.56$, $p(\text{one-sided})=0.058$).

To assess whether approachers exhibited stronger HBVab change or regulation effects compared to avoiders, we conducted an independent t-test between groups with post-vaccination HBVab change, and VTA upregulation effects as the dependent variable. Analyses did not find significant differences between groups (HBVab change: $t(66)=-0.38$, $p=0.701$; VTA upregulation effects: $t(55)=0.47$, $p=0.637$).

Overall, these correlational analyses show that neither the specific VTA upregulation effect, nor the post-vaccination HBVab change, and thus the correlation between them, can be attributed to differences in neurobehavioural motivational tendencies and capacities as measured by these designated tasks and questionnaires.

**Immunological and fMRI-NF pilot studies**

Between January and December 2019 (prior to the initiation of the main trial), we conducted pilot studies to characterize the nature of the immune response to HBV vaccination in our target population, and to test the efficacy of our NF protocol in yielding robust reward-ML activations.

Thirteen participants were recruited for the pilot studies. To characterize the immunological response to HBV vaccination regardless of NF training, three participants underwent blood tests and were vaccinated in similar time intervals to those of the no-NF study group in our main trial (two baseline measurements before vaccination, and three blood tests collected three, fourteen, and twenty-eight days following vaccination). Four additional participants underwent reward-ML fMRI-NF training, HBV vaccination, and immunological assessments to compare the former effects with the downstream immunological effects of reward-ML NF. Group allocation was not randomized, blinded, or pre-registered prospectively like the main trial. In addition to the four reward-ML NF participants, six additional subjects participated in reward-ML fMRI-NF training sessions without HBV vaccination and immunological assessments, resulting in a total of 10 participants for the estimation of reward-ML activations during NF.

Immunological analyses (**Figure S5 a**) demonstrated our ability to detect the increase in HBV antibody levels 14 and 28 days following vaccination (pre and post-vaccination measures were calculated like in the main trial). We also detected a large variability in the post-vaccination response, which supported our decision to collect a relatively large number of participants per group to reveal potential immunological effects. Critically, we found that participants who were vaccinated against HBV approximately 10 years prior to their recruitment can show high baseline HBV antibody levels, which led us to apply a more strict exclusion criterion than the pre-registered one with respect to previous vaccination history, i.e., to exclude subjects that received any prior HBV vaccination other than during infancy (which applied to most of our cohort; described in the main text).

Our fMRI analysis (**Figure S5 b**) validated our four-session NF training length, demonstrating increasingly better regulation effects that peaked at the fourth NF session (In **Figure S5 b** we plot the “Rest” and “Regulate” conditions separately to provide more information about the nature of the regulation effects, which are proportional to the differences in reward-ML activity (y axis) between the two conditions.

### Figure S5: Pilot study results

#### a. Immunological assessments

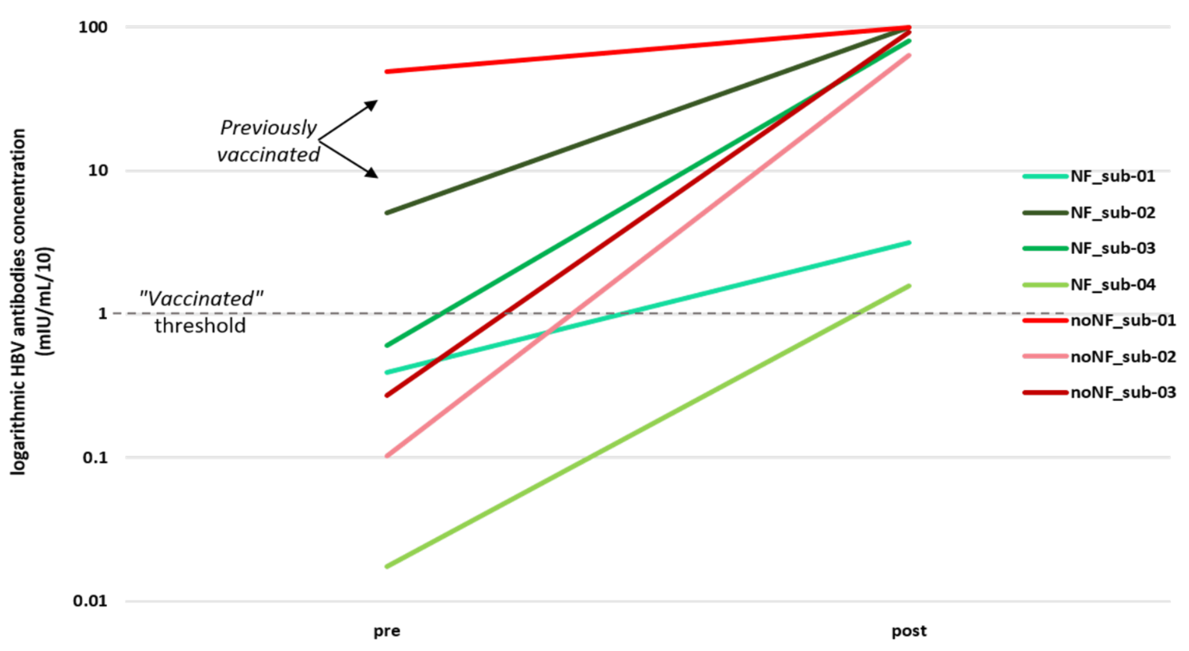


#### b. reward-ML activations during fMRI-NF training

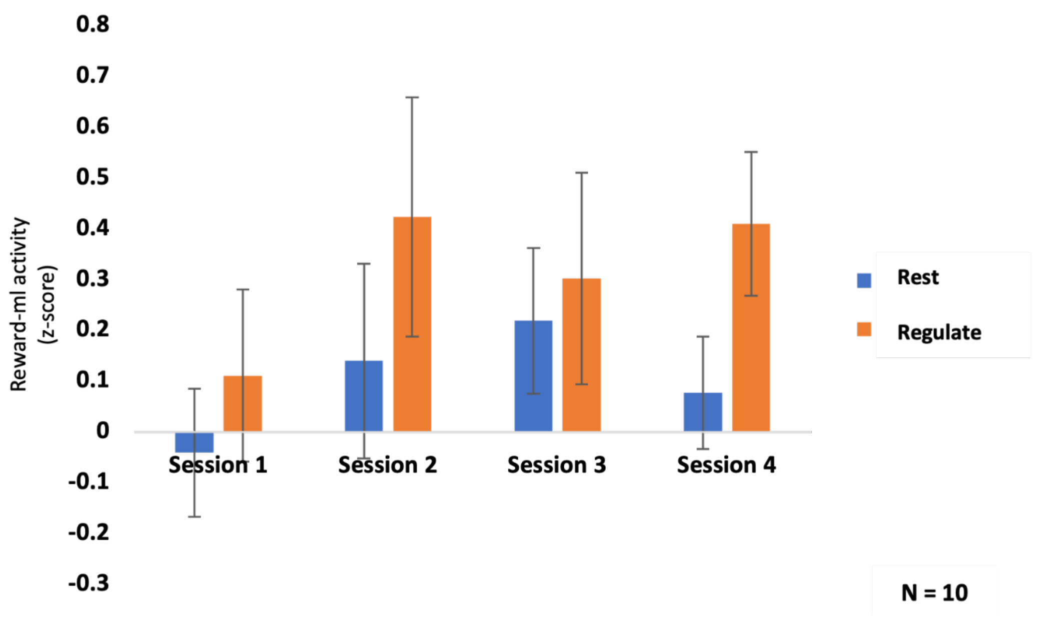
